# Embedded deep-learning based sample-to-answer device for on-site malaria diagnosis

**DOI:** 10.1101/2023.12.10.23299182

**Authors:** Chae Yun Bae, Young Min Shin, Mijin Kim, Younghoon Song, Hong Jong Lee, Kyung Hwan Kim, Hong Woo Lee, Yong Jun Kim, Creto Kanyemba, Douglas K Lungu, Byeong-il Kang, Seunghee Han, Hans-Peter Beck, Shin-Hyeong Cho, Bo Mee Woo, Chan Yang Lim, Kyung-Hak Choi

## Abstract

Improvements in digital microscopy are critical for the development of a malaria diagnosis method that is accurate at the cellular level and exhibits satisfactory clinical performance. Digital microscopy can be enhanced by improving deep learning algorithms and achieving consistent staining results. In this study, a novel miLab^TM^ device incorporating the solid hydrogel staining method was proposed for consistent blood film preparation, eliminating the use of complex equipment and liquid reagent maintenance. By leveraging deformable staining patches, miLab^TM^ ensures consistent, high-quality, and reproducible blood films across various hematocrits. Embedded-deep-learning-enabled miLab^TM^ was used to detect and classify malarial parasites from the autofocused images of stained blood cells by using an internal optical system. The results of this method were consistent with manual microscopy images. This method not only minimizes human error but also facilitates remote assistance and review by experts through digital image transmission. This method can set new paradigm for on-site malaria diagnosis. The miLab^TM^ algorithm for malaria detection achieved a total accuracy of 98.86% for infected red blood cell (RBCs) classification. Clinical validation performed in Malawi demonstrated an overall percent agreement of 92.21%. Thus, miLab^TM^ can become a reliable and efficient tool for decentralized malaria diagnosis.

## Introduction

With over 249 million reported cases and 608,000 casualties as of 2022, malaria is a global public health concern (Geneva: World Health Organization, 2023). The majority of malaria cases (exceeding 99%) are concentrated in low- and middle-income nations, with sub-Saharan Africa accounting for 95% of cases. Despite cost-effective rapid diagnostic tests (RDTs) and precise polymerase chain reaction (PCR), manual microscopy coupled with visual inspection by highly trained experts is widely used for malaria diagnosis because of its robustness and limited false positives and false negatives (Beck, 2022; Fitri et al., 2022). However, smearing and staining quality as well as the expertise of microscopists who read smeared blood slides considerably affect the quality of microscopy-based diagnosis. Malaria eradication in malaria-endemic countries is hindered by the lack adequate healthcare facilities, reagents, trained professionals, vector control, and surveillance systems (Oduola et al., 2018; Sori et al., 2018; Gaston and Ramroop, 2020).

To overcome the challenges of manual microscopic examinations in malaria diagnosis, researchers have proposed digital microscopy and computational image analysis algorithms (Mody et al., 2006). Diagnosis based on digital images can reduce human labor, aid local healthcare workers, and enable experienced experts in remote locations to review microscopic results. Advances in image analysis techniques based on deep learning (Krizhevsky et al., 2012) have rendered this approach more affordable because of its higher accuracy than that of traditional machine learning algorithms (Liang et al., 2016; Gopakumar et al., 2018; Rajaraman et al., 2019; Zhao et al., 2020; Li et al., 2021; Meng et al., 2022; Madhu et al., 2023). However, these studies have only conducted cell-level evaluations on datasets, without clinical tests (Liang et al., 2016; Gopakumar et al., 2018; Rajaraman et al., 2019; Molina et al., 2020; Zhao et al., 2020; Li et al., 2021; Meng et al., 2022; Madhu et al., 2023). Although studies have reported patient-level malaria diagnosis using image analysis in clinical settings, the accuracy of these methods is limited (Yoon et al., 2019; Das et al., 2022). Despite computer algorithms exhibiting greater consistency than that of manual readers, slide quality considerably affects the performance of these computer algorithms (Das et al., 2022). Therefore, computer algorithms should be scaled up with more datasets to overcome their dependency on blood-film preparation quality and ensure adaptability to the variability of slides.

Traditional machine learning algorithms tend to saturate with the increase in the amount of training data. By contrast, deep learning algorithms can learn from larger datasets to train larger models to attain improved accuracy. For example, in 2012, the best model for image classification had 62 million trainable parameters (Krizhevsky et al., 2012), whereas in 2022, the number of parameters increased to 2,44 trillion in 2022 (Wortsman et al., 2023). The required computational resources should be increased to accommodate larger models. Cloud computing or server-level computing is generally used to run current state-of-the-art models. However, in many malaria-endemic countries, using large, extensive, deep models that require powerful computing capabilities or a consistently stable Internet connection is not feasible. By contrast, embedding an efficient deep learning model into a portable device can be prove useful in real clinical environments. Furthermore, consistent blood film preparation is essential to achieve high accuracy by reducing image variability.

In this study, we introduced miLab^TM^, an embedded-deep-learning-based sample-to-answer device capable of automated blood film preparation and autofocused imaging using digital microscopy for on-site malaria diagnostics (Figure 1A). We provided a quick, inexpensive, and environment-friendly method to stain smears with dyes by using deformable staining patches. The automated preparation provided by miLab^TM^ reduces differences in staining methods between technicians or institutions, resulting in consistent and high-quality preparation results for blood samples over a broad range of hematocrits. (Choi et al., 2021; Bae et al., 2023). The embedded deep learning algorithm was used to analyze malaria-suspected morphology from the blood cells stained in this device, and the autofocused digital images were captured by the optical system of the same device. The proposed miLab^TM^ device can perform continuous autofocus imaging, providing the same effect as continuous field-of-view (FoV) readings applied when examined under a microscope. Digital microscopy allows scanning of more than 200,000 RBCs within 7–10 min without errors, which is more than the number of RBCs recommended by WHO guidelines. Therefore, digital microscopy is beneficial for samples from patients with low parasitemia (in cases of low infection). Therefore, this innovative device allows on-site users to analyze digital image data immediately and obtain immediate diagnostic results without high-performance computing. Simultaneously, digital images can be sent to experts from remote locations to assist with diagnosis (Figure 1B). On-site users can immediately check the suspected morphology of malaria-infected cells through the screen mounted on the device (Figure 1C), and other experts can access the same digital images on web-based software to review the suspected morphologies and diagnostic results (Figure 1D). The embedded deep-learning-based on-site malaria diagnostic platform not only provides similar results as in-person microscopy examination in the laboratory but also enables remote diagnosis.

**Figure 1.**
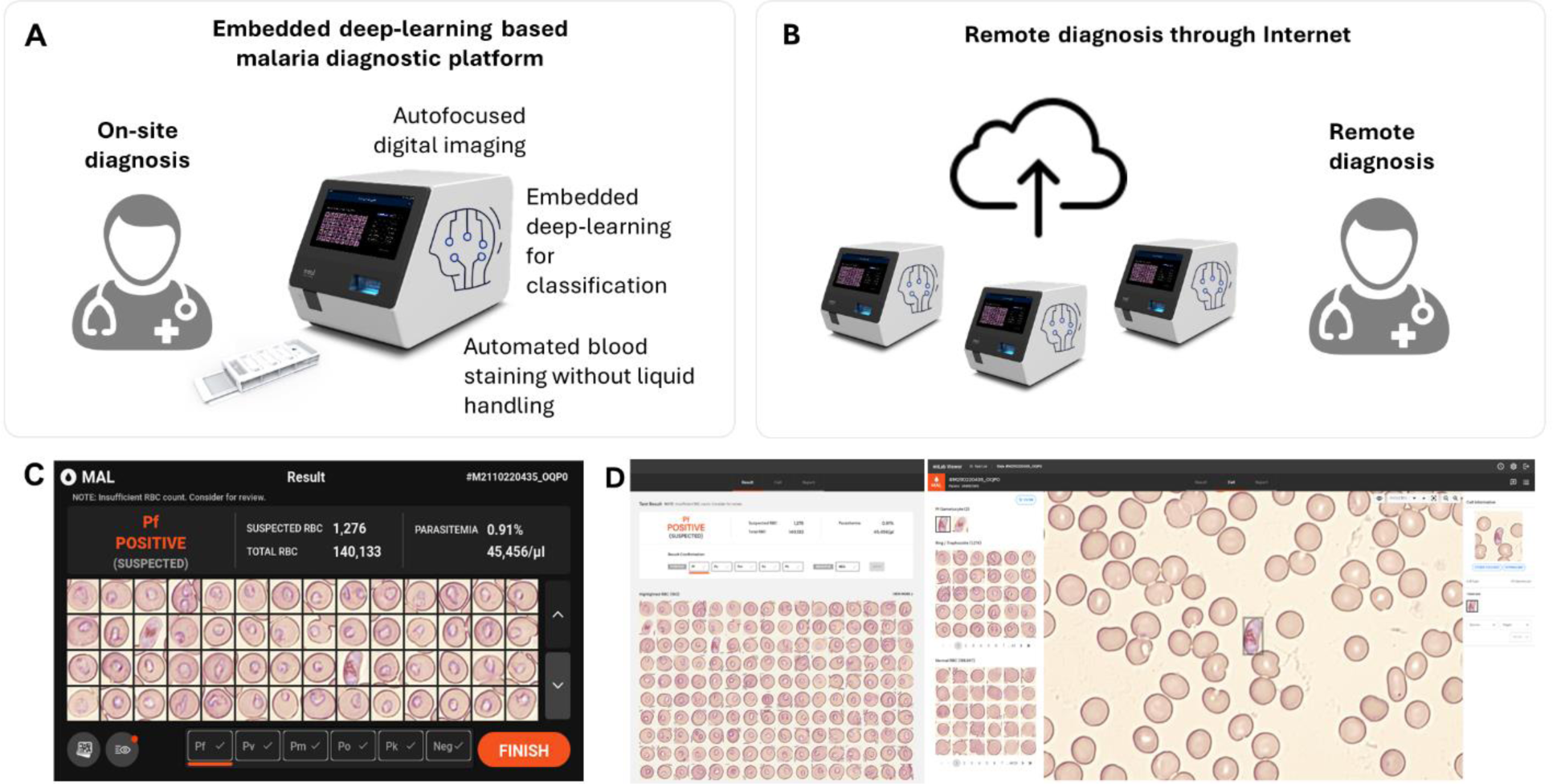
Schematic of the embedded deep-learning based on-site malaria diagnosis. (A) The miLab^TM^ device not only automates the process (automated blood staining without liquid handling and autofocused digital images) of malaria diagnosis through microscopic analysis but also incorporates deep learning algorithm directly into the device for on-site review. (B) A web-based software allows experts to access the digital images for remotely reviewing the result through the internet. (C) Photograph of the result page in miLab^TM^ for *P.falciparum* positive patient specimens. Users can review and confirm the results in the miLab^TM^ for sample-to-answer, on-site malaria diagnosis. (D) Photograph of the screen shot of the result page from the same patient specimens on the web-based software, accessing remotely digital images and raw data from miLab^TM^. Other experts can remotely review and confirm the same results from miLab^TM^.

## Materials and Methods

### miLab^TM^ Platform

The miLabTM platform was designed to provide fully automated microscopic analysis, which is the standard for malaria diagnostics. Each cartridge (40 mm (width) × 92 mm (length) × 15 mm (height). w: width, l: length, and h: height) of the miLabTM device (212 mm (w) × 390 mm (l) × 244 mm (h). w: width, l: length, h: height) consisted of a spreader film for preparing the blood film and three types of staining patches, facilitating consistent RBCs smearing and staining (Supplementary Figure S1). The cells can be smeared and stained by simply moving 5 μL of blood-loaded cartridge without any controlling the aqueous solution. Thus, automated processes for blood films can be designed to stain the morphology of *Plasmodium* effectively and appropriately on-site. We used a solid staining method using hydrogels to eliminate the need to control complex equipment or maintain the liquid reagents (Choi et al., 2021). In this approach, the hydrogel is brought into contact with appropriately smeared and fixed cell surfaces, allowing the dye to efficiently stain the cells. Depending on the type of dye used, the dye from the hydrogel was applied to the smear in a short time, under 1 minute, depending on the type of dye used (Bae et al., 2023). Furthermore, hydrogels without dye can absorb any remaining dye on the cell surface and attain suitable staining quality depending on the pH of the buffer in the hydrogel, which is similar to the role of the conventional buffer solution in blood cell staining (Oktiyani et al., 2023). After automated blood-film preparation, the optical system of the device captured the digital images of the stained blood cells. At least 200,000 RBCs and up to 500,000 RBCs were scanned in continuously obtained digital images from each blood film. The optical system was designed to capture multifocal images from the blood film by considering the size and location of the stained parasites within the RBCs.

Subsequently, a machine learning algorithm was applied to analyze the captured images and detect malarial parasites in the blood. To incorporate the machine learning model into the limited resources of the embedded hardware, all neural network architectures were designed to reduce computational complexity. To identify parasites that are rarely present in RBCs, we devised a two-step image analysis algorithm, including RBC detection and subsequent classification. The subimages were extracted from the image to obtain one RBC for each event (1st step: detection). Each of the cropped subimages was then tested for the presence of malarial parasite (2nd step: classification). In the two-step algorithm, images are divided into meaningful units and each unit is analyzed comprehensively, instead of finding scarce malarial parasites directly in numerous images.

In the detection module of miLabTM, a semantic segmentation algorithm is used to detect each RBC in the images. Because our target images were full of RBCs, we adopted a semantic segmentation algorithm instead of a general object detector to achieve compactness and efficiency (Ronneberger et al., 2015; Tran et al., 2019; K.T. et al., 2022). The model was specifically designed to produce two outputs, namely objectness and contours, as segmentation mask images. After running the semantic segmentation algorithm, the contour image is subtracted from the objectness image to separate the aggregated cells. The cell locations were determined by performing the connected component analysis from the resulting mask (Supplementary Figure S3A). Detected blood cells were cropped and passed through the classification module.

In the classification module, we used the fact that uninfected cells were the dominant component of a large number of detected cells. Because passing all cells through the entire classification procedure is inefficient, we designed the classification module as two cascaded classifiers to reduce the computational burden imposed by the skewed ratio of parasite-positive to parasite-negative cells (Supplementary Figure S3B). The first classifier consists of only three convolutional layers for the maximum speed and screens for obviously “clean” uninfected cells. If a cell was clearly classified as uninfected, then it was not passed on to the second classifier. This prior “screening” classifier considerably reduces the number of cells delivered to the subsequent classifier. The second “main” classifier performs an in-depth examination of only those cells deemed by the first classifier to be possibly infected. The second classifier is based on the ResNet (He et al., 2016) architecture and is equipped with a convolutional block attention scheme (Woo et al., 2018). Similar to our segmentation network, the main classifier has a multi-task learning framework with two output branches: one branch that predicts the presence of malaria (infection branch) and the other branch that estimates its developmental stage (stage branch). In our verification study, only 17.8% of the RBCs passed the screening classifier and were delivered to the main classifier, whereas the conventional monolithic classifier examined all RBCs (Supplementary Figure S4). The classifier was trained to be robust to distracting elements, such as white blood cells (WBCs), which are categorized as malaria-negative. The current AI of miLabTM can detect malaria parasite-infected RBCs in the operator-assigned number (default: 200,000) of RBCs and classify developmental stages based on their morphology: *Plasmodium falciparum* and *Plasmodium vivax* ring stage, *P. falciparum* late stage (i.e., gametocyte), *P. vivax* trophozoite stage, and *P. vivax* late stage (i.e., schizont and gametocyte).

The algorithm of the device provides flexibility in the tradeoff between sensitivity and specificity. The output score of the infection branch was thresholded to determine whether the cells were positive or negative. Lowering the threshold yields higher sensitivity and raising the threshold provides higher specificity. Furthermore, patient-level recommendations are provided based on the number of suspected cells on a slide. In this clinical evaluation, we considered a slide with more than one positive RBC as malaria-positive..

Analysis of the blood film in the miLab^TM^

Whole blood samples were collected for research purposes, and the study was approved by the Institutional Review Boards (P01-202003-31-007 and GCL-2020-1011-01) of the Korea National Institute for Bioethics Policy (Seoul, Korea) and GCLabs (Yongin, Korea). The hematocrit of the blood sample was measured using a hematocrit-measuring instrument to analyze the blood film (Boditech Med Inc., FPRR005). The blood film was analyzed based on the number of RBCs and color value of the prepared stained RBCs. Images of stained RBCs prepared using the specimens were captured in the device, and the field of views (FoVs), including stained RBCs, were acquired. The number of RBCs was analyzed using a verified image analysis tool. Color values (red, green, and blue (RGB)) were obtained from the pixels of the segmented RBCs using a verified, self-made image analysis tool. To examine the morphological features of *Plasmodium*, four types of typical stages and species (rings of early trophozoites, late trophozoites from *P. vivax*, and gametocytes from *P. falciparum* and *P. vivax*) were collected. To compare the staining quality of *Plasmodium* in miLab^TM^, a manual Giemsa slide was prepared with the same blood samples and observed under a microscope (CX33 with a 100x objective lens, Olympus). The same morphology of *Plasmodium* in the blood film prepared from miLab^TM^ was observed using the same device and under a microscope (CX33 with a 50x objective lens, Olympus) to compare the image quality of miLab^TM^.

### System verification of miLab^TM^

To evaluate the reproducibility of blood films smeared using miLab^TM^, a verified, self-made image analysis tool was used to determine the number of segmented RBCs in each FoV. The mean RBC counts from 50 FoVs of 20 replicate slides for seven specimens were used to examine the precision of the blood smear. To evaluate the reproducibility of blood film staining by miLab^TM^, a verified, a similar self-made image analysis tool was used to determine the mean RGB value of segmented RBCs from 200 FoVs. Two hundred FoVs per specimen were used to examine the color of the specimen by calculating the mean, standard deviation, and coefficient of variance.

A receiver operating characteristic (ROC) curve was drawn using the binary classification result of 4,005 normal RBCs and 7,713 malaria positive cells. The classification performance at the cellular level for *Plasmodium* (ring and gametocytes) was represented using a confusion matrix (2 × 2), where the maximum accuracy was obtained. The detection rate of *Plasmodium* (ring and gametocyte) for the entire embedded deep-learning algorithm was verified using the *Plasmodium* ratio among the total number of RBCs per FoV obtained from the blood film in miLab^TM^. In total, 3,000 FoVs from 15 malaria-positive clinical specimens (200 FoVs each) from the blood film in miLab^TM^ were labeled by an experienced microscopist. A total of 3,290 *Plasmodium* (ring, gametocyte) were confirmed in 1,751 FoVs. The detection rate of *Plasmodium*, determined by the embedded deep-learning algorithm in miLab^TM^ (test group), was compared and fitted to that observed by the human eye on the same images (control group).

### Clinical evaluation

Clinical samples (n = 555) analyzed using the two miLab^TM^ devices were collected from April 2022 to November 2022 and used in the clinical validation study approved by the Institutional Review Board (IRB00003905) of the National Health Sciences Research Committee (Ministry of Health, Malawi). After explaining the purpose of the study, procedure, possible benefits, risks, and rights of the participants, all participants were first requested to sign informed consent forms. The results obtained by miLab^TM^ were compared to those of microscopic examination by an experienced local microscopist in Malawi and alongside with RDT (CareStart™ Malaria Pf (HRP2) Ag RDT, AccessBio, NJ). Blood samples (∼250 µL) were collected into blood capillary tubes by a finger prick through a sterile lancet and stored in an anticoagulant tube with ethylenediaminetetraacetic acid (EDTA). Five microliters of collected blood was loaded onto the miLab^TM^ cartridge and five microliters of the collected blood was used to prepare a thick and a thin blood film each for microscopic examination. The blood film was stained with a mixture of eosin and methylene blue using Giemsa staining. Local microscopists examined the Giemsa-stained slides using an Olympus CX33 microscope according to the standard microscopy methods of the World Health Organization (WHO & UNICEF/UNDP/World Bank/WHO Special Programme for Research and Training in Tropical Diseases, 2015). Parasitemia determined using miLab^TM^ (test group) was compared and fitted linearly to that measured from the thick film of the manual Giemsa slide under microscopic examination by a local microscopist (control group).

### Statistical analysis

GraphPad Prism (Ver.7, GraphPad Inc., San Diego, CA, USA) was used to perform all statistical analyses. To evaluate the linear correlation for the detection rate in the system verification and parasitemia in clinical evaluation, linear least squares analysis was performed at the 95% confidence interval of each variable, and the Pearson correlation coefficient (*r*) was calculated. Differences between the test and control groups were examined using Student’s unpaired t-test. A two-sided test, and the results were considered statistically significant at *p* < 0.05.

## Results

### Characterization of the blood film in the miLab^TM^

To address the unique characteristics of malaria patients with lower RBC counts, we implemented a two-speed smearing process on a spreader with a consistent angle, screening an adequate number (at least more than 200 RBCs per FoV) of RBCs in low hematocrit samples, ranging from 20% to 35%. This method provided two zones for detecting appropriate RBCs, depending on the hematocrit of the samples (Figure 2A). When we investigated the correlation between the average RBC counts per FoV and the hematocrit from the 37 clinical specimens, zones A and B revealed a linear correlation between RBC counts and hematocrit (Figure 2B, Supplementary Figure S5). The RBC counts of samples with low (< 30%) and middle/high hematocrit (> 30%) were selected from zones B and A, respectively. To increase the efficiency of RBC screening, RBCs from low-hematocrit samples can be screened in zone B (average 200–400 RBC counts per FoV) instead of zone A (average 100–300 RBC counts per FoV).

**Figure 2.**
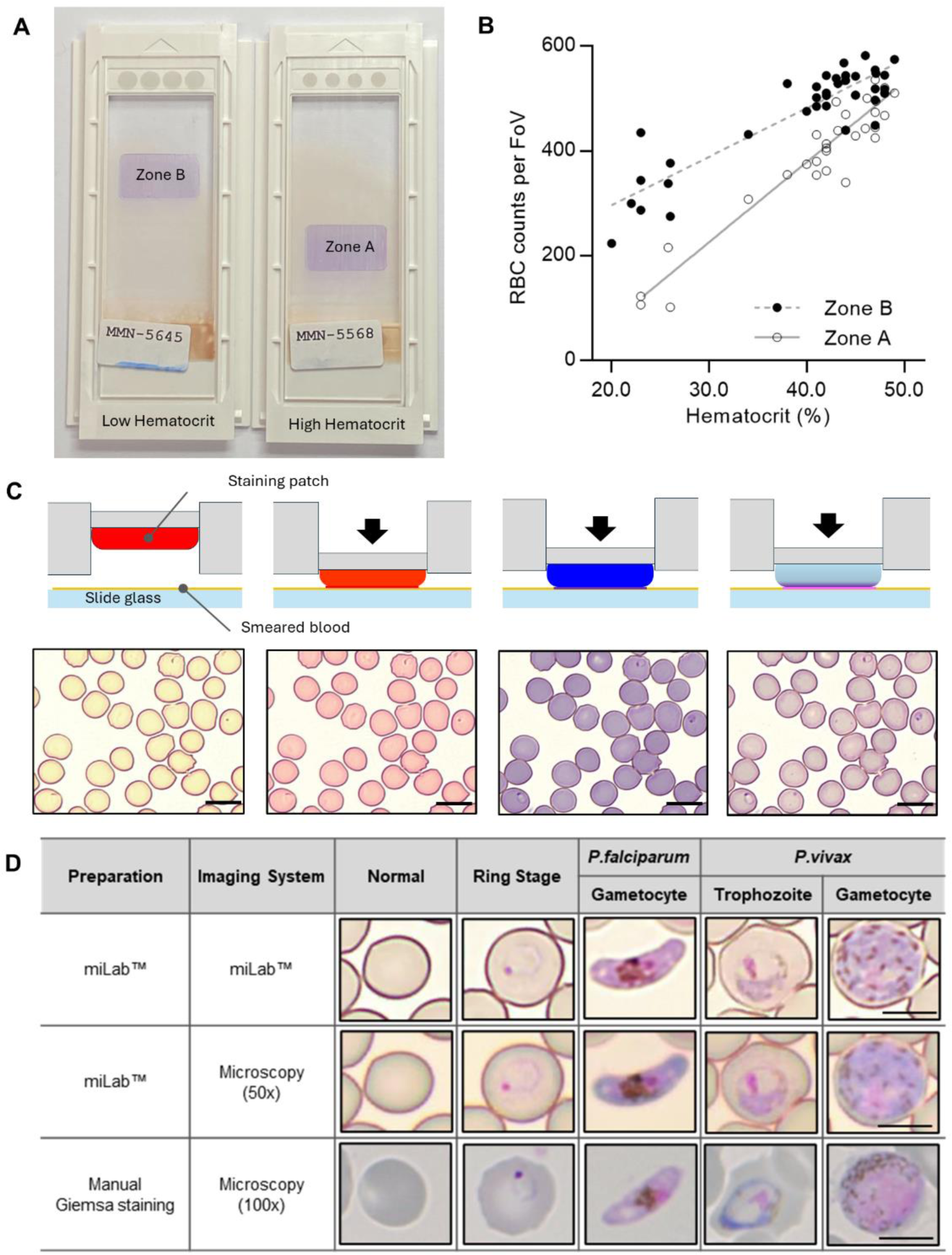
Characterization of the blood film in the miLab^TM^. (A) Photograph of the prepared blood films from the miLab^TM^ using a patient specimens with low hematocrit and high hematocrit from Malawi. Low hematocrit samples to be read in Zone B instead of Zone A, where high hematocrit samples were typically read. The miLab^TM^ device automatically detects an appropriate area to observe RBCs in a monolayer. (B) Correlation of average RBC counts per FoV depending on the hematocrit of the clinical specimens (n = 37) was shown with open dots (Zone A) and close dots (Zone B). (C) Schematic of blood staining using three distinct staining patches in the cartridge and pictures of stained blood cells with *Plasmodium*-infected RBCs (black arrow) from each steps of staining procedure. The scale bars = 10 μm. (D) Comparison of microscopic cell image with the miLab^TM^ blood film acquired from miLab^TM^, 50x olympus microscopy with miLab^TM^ blood film, and 100x microscopy with conventional Giemsa slides. The scale bars = 5 μm.

Depending on the types of staining patches that included various types of Romanowsky stains, such as Eosin, Methylene blue, and Azure B, the color of RBCs and the morphology of parasites were observed at each staining step (Figure 2C). The transparent patch was used for absorbing the excess dye left on the slide and optimizing the stained colors of the cells, whereas the dye-containing patches were used to deliver dyes to the cells (Choi et al., 2021; Bae et al., 2023). To confirm that the morphological characteristics of *Plasmodium* were revealed in the blood film prepared in miLab^TM^, four types of typical stages and species (rings for early trophozoites, late trophozoites from *P. vivax*, and gametocytes from *P. falciparum* and *P. vivax*) were collected from patient samples (Figure 2D). The stained blood film provided a clear morphology of *Plasmodium* at each stage, similar to conventional microscopic examination using Giemsa staining for 100x microscopy images. In particular, when comparing the images of *Plasmodium* morphologies stained by the cartridge with manually focused images at the same resolution using a 50x microscope, the distinctions in morphology and species were discernible, even in the auto-focused images within the device. Both automatically stained cells and their autofocused digital images not only allow the embedded deep-learning algorithm to detect malaria on-site but also enable experts to distinguish between types and stages of malaria through the result screen in the device or viewer at remote locations.

### System verification of the miLab^TM^

To ensure an accurate diagnosis, digital images that demonstrate reproducible automated preparation processes and effectively depict the morphology of *Plasmodium* are crucial. Therefore, when performing automated preparation processes in miLab^TM^, the blood film was verified by the consistency of RBC counts for smearing and the color difference for staining. The screening of RBCs performed on various hematocrit samples, including 20%–50%, resulted in a coefficient of variation (CV%) of less than 10%, even in 20 replicates of seven clinical specimens (Figure 3A). Because three-color values (red, green, and blue [RGB] levels) are the most common color spaces for segmenting parasites and RBCs from thin blood films (Fong Amaris et al., 2022), each color value was extracted from segmented RBCs to quantitatively analyze the appropriate blood smear and staining. Thus, hydrogel staining precisely controls the color of *Plasmodium* (ring, gametocyte) and performs blood film preparation efficiently and reproducibly. Figure 3B displays the color values of the stained images. The %CV of the three-color values over 4,000 FoVs was less than 5% and was maintained across 200 FoVs for 20 specimens (Supplementary Figure S6).

**Figure 3.**
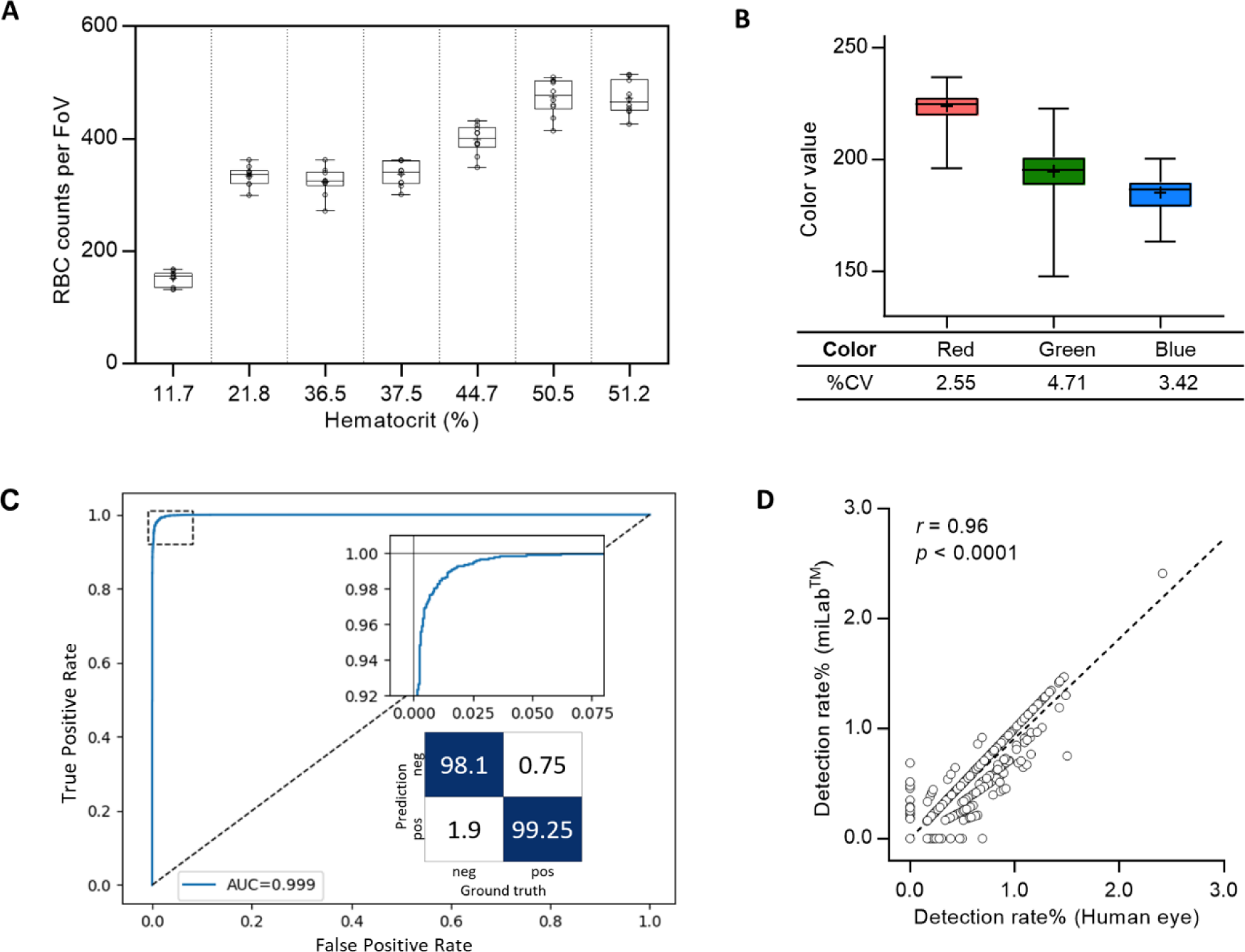
System verification of the miLab^TM^ device. (A) Reproducibility of blood smear was represented with the box plot using RBC counts per FoV in seven clinical specimens (n = 20). Average RBC counts per FoV were demonstrated with low, middle, and high hematocrits. The RBC counts of the samples with the low (< 30%) and the middle/high hematocrits (> 30%) were selected from Zones B and A, respectively. (B) Reproducibility of blood staining was represented with the box plot using the red, green, and blue color value, which was obtained from the stained RBCs in FoVs of clinical specimens. The RGB color values of each RBC was conserved across FoVs (n = 4,000). (C) Cellular level classification performance for *Plasmodium* (ring, gametocytes) was represented with the ROC curve. The area under the curve (AUC) was 0.999 with 95% confidence interval in the range of 0.9986–0.9994. The confusion matrix was calculated at the optimal point where maximum accuracy was obtained. (D) Correlation of the detection rate of malaria positives (ring, gametocytes) between the deep learning algorithm (test group: miLab^TM^) and naked eyes (control group: Microscopy) with the Pearson’s correlation coefficient (r) of 0.96 (p < 0.0001, n=3,000).

The ROC curve in Figure 3C displays the cellular-level performance of *Plasmodium* binary classification. The area under the curve (AUC) was 0.999, indicating that the proposed classifier was highly accurate. The magnified graph displays the trade-off relationship between the true positive and false positive rates. We selected an optimal point on the curve that achieved the maximum accuracy and calculated the sensitivity, specificity, and accuracy at that point to be 99.25%, 98.1%, and 98.86% (95% CI: 98.65–99.04%), respectively. Infected RBC detection performance was verified using blood films prepared from 15 malaria-positive clinical specimens. We randomly selected 200 FoVs for each blood film and compared the proportion of infected RBCs from miLab^TM^ to microscopic results obtained visually. The classifier infection branch applied an empirically determined threshold value to the output. A cell is classified as malaria-positive if its output value surpasses a specified threshold. An excellent correlation existed between the results from both methods, with a Pearson’s correlation coefficient (*r*) of 0.96 (*p* < 0.0001; Figure 3D).

### Evaluation of Clinical Performance in Malawi

The clinical performance of miLab^TM^ was evaluated using 555 patients who were selected from patients with fever and visited the clinical study site, Mzuzu Health Center in Malawi. The predominant species was *P. falciparum* (Gaston and Ramroop, 2020; U.S. President’s Malaria Initiative Malawi Malaria Operational Plan FY, 2020). Clinical information of the patients is summarized in Supplementary Tables S1 and S2. Figure 4A depicts the study design for the clinical validation of miLab^TM^ based on the approved study protocol. The miLab^TM^ results were compared in concordance with those of manual microscopy (local and expert microscopists) and RDT as reference tests. On analysis of 488 clinical specimens by miLab^TM^ after excluding 67 samples with discrepancies between reference tests, an overall percent agreement (OPA) of 92.21% (95% confidence interval (CI); 89.48%–94.43%), positive percent agreement (PPA) of 95.15% (95% CI; 89.03%–98.41%), and negative percent agreement (NPA) of 91.43% (95% CI; 88.17%–94.03%) was observed (Figure 4B).

**Figure 4.**
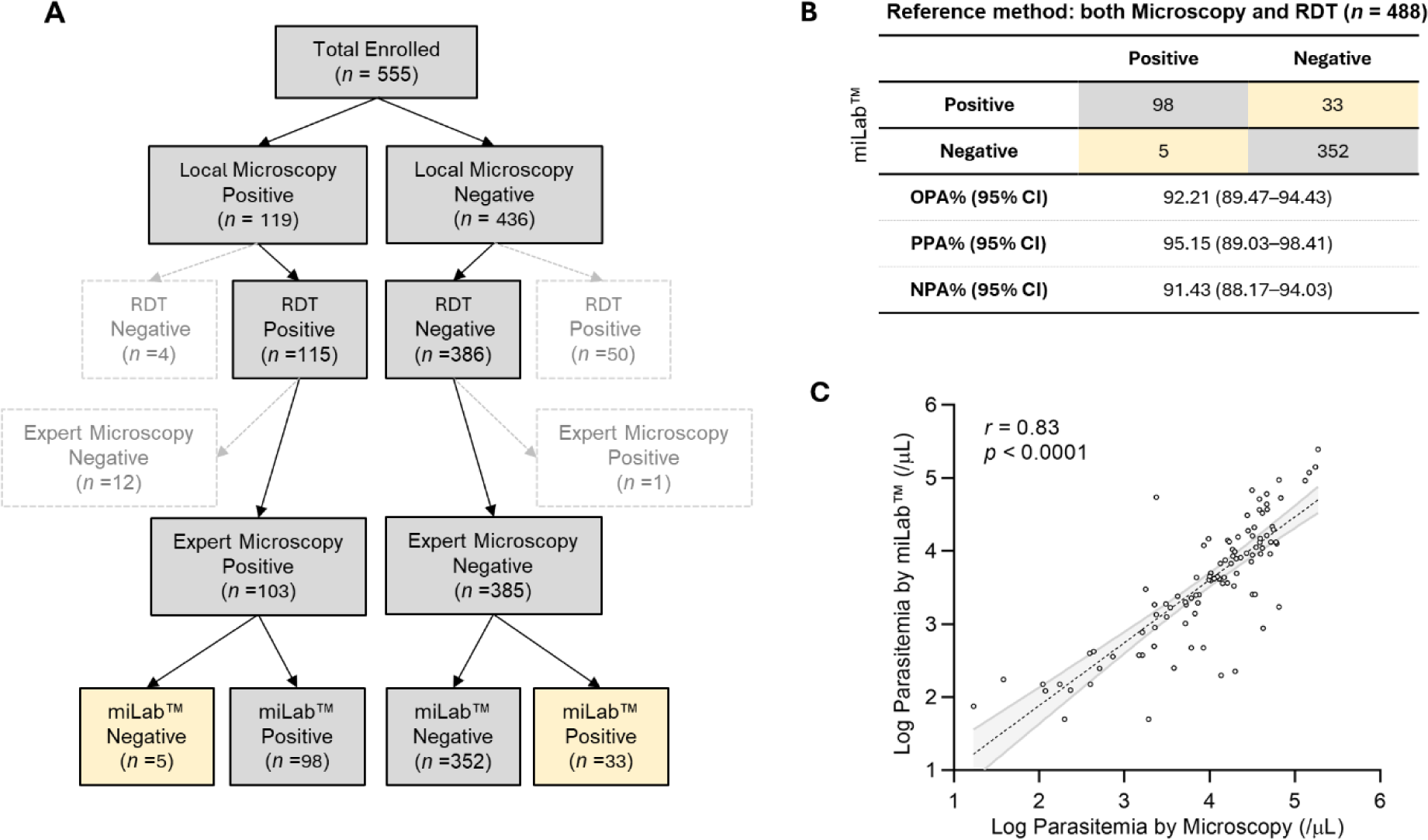
Clinical validation of miLab^TM^ in Malawi. (A) Design for a clinical study. A total of 555 clinical specimens were enrolled and subjected to microscopy and analys^is^ by miLab^TM^ for comparison with the reference tests (both local microscopy examination and RDT). Yellow cells indicate samples discordant with the reference test. (B) Agreemen^t o^f analysis by miLab^TM^ with the reference tests (microscopy and RDT). Based on the concordance of microscopy and RDT, overall percent agreement (OPA), positive percent agreement (PPA), and negative percent agreement (NPA) were 92.21%, 95.15%, and 91.43% respectively. (C) Correlation of the parasitemia level between microscopy and the miLab^TM^ on a logarithmic scale. The Pearson’s correlation coefficient (*r*) is 0.8259 (95% CI: 0.7518 to 0.8794).

Figure 4C displays the correlation of parasitemia between manual microscopic examination by a local microscopist and miLab^TM^. The mean parasitemia of positive samples is approximately 26,000 (parasites per µL) from the miLab^TM^ and approximately 22,000 (parasites per µL) from the local microscopist. Because miLab^TM^ quantifies parasitemia assuming 5,000,000 RBCs per microliter of the blood, parasitemia in patients with abnormal RBCs or out-of-range WBCs differs from quantification based on 8,000 WBCs per microliter of blood through the conventional microscopy examination. Nevertheless, the Pearson’s correlation coefficient (0.8259, 95% CI: 0.7518–0.8794) of parasitemia determined by miLab^TM^ exhibited excellent consistency with the quantification results obtained from Giemsa slides by the microscopist. Based on the results of the parasitemia level, 200 clinical samples (100 positive, 100 negative) were randomly selected and the limit of detection (LOD) for the point at which a positive result is more than 95% probable by using probit regression is approximately 31 parasites per μL.

## Discussions

On-site sample-to-answer malaria diagnosis in miLab^TM^ enables blood film preparation for embedded deep learning-based malaria detection using digital microscopy images. In miLab^TM^, hydrogel-stained patches are applied to blood film generation in a highly reproducible manner. In this on-site diagnostic platform with an A4 paper-sized footprint, ethanol-based fixation is applied to avoid the use of methanol and staining patches to reduce liquid waste without maintaining reagents for user safety and convenience. Thus, a sophisticated, well-equipped central laboratory is not required for on-site diagnosis. The miLab^TM^ device functions as a stand-alone unit that can be used in resource-limited environments. System validation revealed excellent reproducibility for blood film preparation (Figure 3, Supplementary Figure S5). The accuracy of deep-learning-based analysis at the cellular level was comparable to that of other research groups (Liang et al., 2016; Shen et al., 2017; Gopakumar et al., 2018; Rajaraman et al., 2019; Zhao et al., 2020; Li et al., 2021). The consistent performance and higher accuracy of miLab^TM^ eliminated the dependency on technicians for manual microscopy-based malaria diagnosis by providing both blood film preparation and automated analysis compared with other products (Yoon et al., 2019; Das et al., 2022).

The clinical performance evaluation of miLab^TM^ using 488 clinical specimens revealed an overall percentage agreement (OPA) of 92.21% before user reviews. Of the 488 samples analyzed, 38 (7.79%) were discordant with the reference tests as false-negatives or false-positives. The miLab^TM^ device displays “review needed” or “suspected” morphology of the parasites on its display to users who wish to review the results either on the device or web-based software. In this clinical validation study, users were required to review the raw data and confirm the diagnostic results for 131 of the 488 patient specimens (including 98 true positives and 33 false positives). To achieve optimal results expected from the device, users can set a cellular-level threshold according to the situation, such as the presence of an expert. Although we attained high cellular-level accuracy, a special strategy is required to provide patient-level recommendations. Because the number of normal RBCs is greater than the number of infected RBCs, setting a cellular level threshold for high specificity can reduce reviewing effort. However, to detect *Plasmodium* in patients with low parasitemia, users should be able to determine patient-level positivity when any cell with a suspected morphology is confirmed on a single slide. Therefore, all suspected cells, even a single cell need to be shown on the Results page. These strategies are included in patient-level recommendations.

The first generation of malaria diagnosis using miLab^TM^ was based on the characteristic morphology of *P. falciparum*. However, the results obtained by a local microscopist confirmed that four patients were infected with *P. vivax* and *P. malariae* in addition to *P. falciparum*. On reviewing the digital images of miLab^TM^, experts observed various distinct morphologies of *Plasmodium* species. The current stage classification performance was examined using 11,718 single-cell images acquired from blood films prepared using miLab^TM^, where the stages were determined independently by expert microscopists. Figure 5B displays the confusion matrix from the verification study for multistage malaria parasite classification. The total accuracy for the classification of infected RBCs was 98.83% (95% CI: 98.62%–99.01%), and that for the classification of *Plasmodium* species and stages was 97.82% (95% CI; 97.53%–98.06%). The miLab^TM^ algorithm revealed excellent performance in malaria detection, with the identification of multiple stages of the parasite’s life cycle in this analytical validation study. If the algorithm is optimized at patient-level estimation, the performance of miLab^TM^ can be improved, allowing the classification or detection of other *Plasmodium* species. A deep learning algorithm trained with other types of infected blood cells would improve the performance of malaria diagnosis by reducing the interference caused by WBCs, platelets, and other types of parasites (e.g., *Trypanosoma cruzi*).

**Figure 5.**
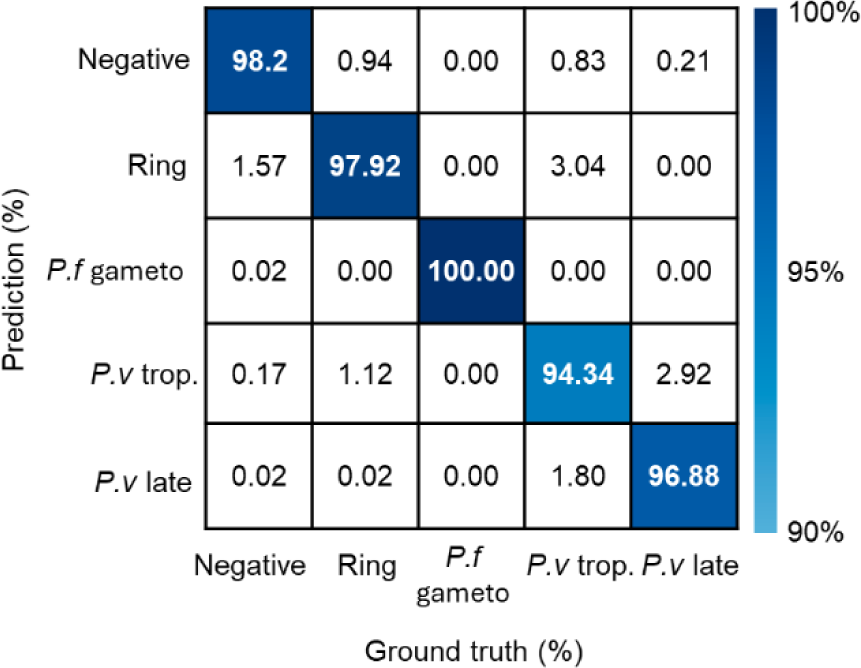
Performance of detecting stages and species in the miLab^TM^. The confusion matrix represented stage and species classification using embedded deep learning algorithm with additional patient-level estimation. (*P.f* gameto; gametocyte, *P.v* trop.; trophozoite, *P.v* late; schizonts or gametocyte).

The miLab^TM^ device can provide on-site, sample-to-answer diagnostics for malaria and enables decentralized patient care in resource-limited settings, especially low- and middle-income countries. In future studies, the miLab^TM^ cartridge can be applied to the morphological detection of WBCs because similar methods derived from Romanowsky-type staining are used for staining for both WBCs and malaria (Supplementary Figure S7A)(Choi et al., 2021). Moreover, by modifying the composition of the hydrogels in the cartridge, miLab^TM^ can prepare various types of sample slides. These slides included cytology slides with Papanicolaou staining (Supplementary Figure S7B) and tissue sections stained with hematoxylin and eosin (H&E) for histopathology (Supplementary Figure S7C) (Chin et al., 2022; Kim et al., 2023). The miLab^TM^ device can be used to diagnose other infections and the detection of intestinal parasites (Supplementary Figure S7D) that require microscopic examination without staining (wet preparations). The use of miLab^TM^ can be extended to the diagnosis of multiple diseases using the external training of deep-learning algorithms on various types of digital images.

## Conclusions

On-site sample-to-answer malaria diagnosis using miLab^TM^ enables blood film preparation for embedded deep learning-based malaria detection using digital microscopy images. The miLab^TM^ algorithm achieved 98.86% detection accuracy of infected RBCs. The clinical validation of miLab^TM^ demonstrated an OPA of 92.21% in Malawi. This on-site malaria diagnostic platform can assist experts in evaluating the suspected morphology of *Plasmodium* in laboratories and remote locations and realize remote diagnosis of malaria, especially in resource-limited settings.

## Data Availability

All data produced in the present study are available upon reasonable request to the authors

## Author Contributions

CYB: Data curation, Formal analysis, Investigation, Methodology, Visualization, Writing - original draft, Writing - review and editing, YMS: Software, Formal analysis, Investigation, Methodology, Visualization, Writing - original draft, Writing - review and editing, MK: Formal analysis, Project administration, Investigation, Methodology, Visualization, Writing - original draft, Writing - review and editing, YS: Formal analysis, Data curation, Visualization, Writing - original draft, Writing - review and editing, HJL: Data curation, Project administration, Software, Writing - review and editing, KHK: Conceptualization, Data curation, Methodology, Software, Writing - review and editing, HWL: Data curation, Project administration, Software, Writing - review and editing, YJK: Data curation, Project administration, Writing - review and editing, CK: Data curation, Writing - review and editing, DKL: Investigation, Supervision, Writing - review and editing, BK: Formal analysis, Visualization, Writing - original draft, SH: Formal analysis, Visualization, Writing - original draft, HPB: Investigation, Supervision, Writing - review and editing, SC: Data curation, Writing - review and editing, BMW: Data curation, Formal analysis, Writing - review and editing, CYL: Conceptualization, Funding, Investigation, Resources, Writing - review and editing, KC: Investigation, Supervision, Validation, Writing - review and editing.

## Acknowledgements

We thank the following associates from Noul Co., Ltd. for their contributions to the development of hardware, software, and the verification of miLab^TM^: H. Lee, S. Moon, J. Cho, Y. Shin, S. Hong, R. Choi, D. Ham, O. Bailo, M. J. Seol, Y. Hong, S. Yun, H. Hwang, E. Hwang, S. K. Beak, and J. Choi. We thank J. Lee, H. Park, and J. Shin for monitoring the IRB project and the Mzuzu Health Centre study team for collecting the clinical data. We thank H. H. Cho for reviewing and editing the manuscript. This This study was supported by a grant from the Korea Health Technology R&D Project through the Korea Health Industry Development Institute (KHIDI), funded by the Ministry of Health and Welfare, Republic of Korea (grant number: HI18C1685).

