## Supplemental Figures for "Embedded deep-learning based sample-to-answer device for on-site malaria diagnosis"

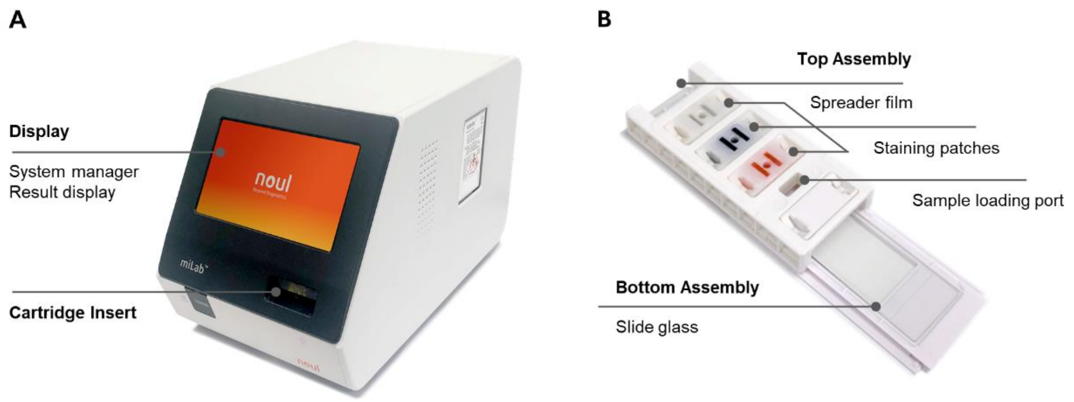


**Supplementary Figure 1. The miLab™ Platform.** (A) The miLab™ device is a fully automated blood film preparation and integrated microscopy image analysis device with embedded deep-learning algorithm. The cartridge is loaded through the cartridge insert port. (B) The miLab™ cartridge prepares a stained blood film from peripheral blood samples and consists of top and bottom assemblies. The top assembly plays three roles; (1) spreader film smears bloods onto the bottom assembly, (2) three staining patches contain solutions for Romanowsky staining; eosin (red), methylene blue/azure B (blue), buffer (transparent). The patches stain the blood cells by sequential stamping. (3) 5 μL of blood are loaded into the cartridge through sample loading port. A standard microscope slide is attached on the bottom assembly.

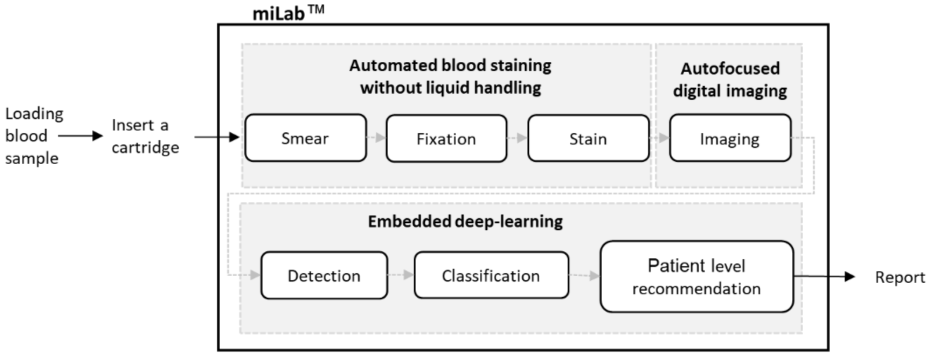


**Supplementary Figure 2. Workflow in the miLab™ platform.** Three procedures in miLab™ included automated blood staining without liquid handling, autofocused digital imaging, and image analyzing using embedded deep-learning algorithm.

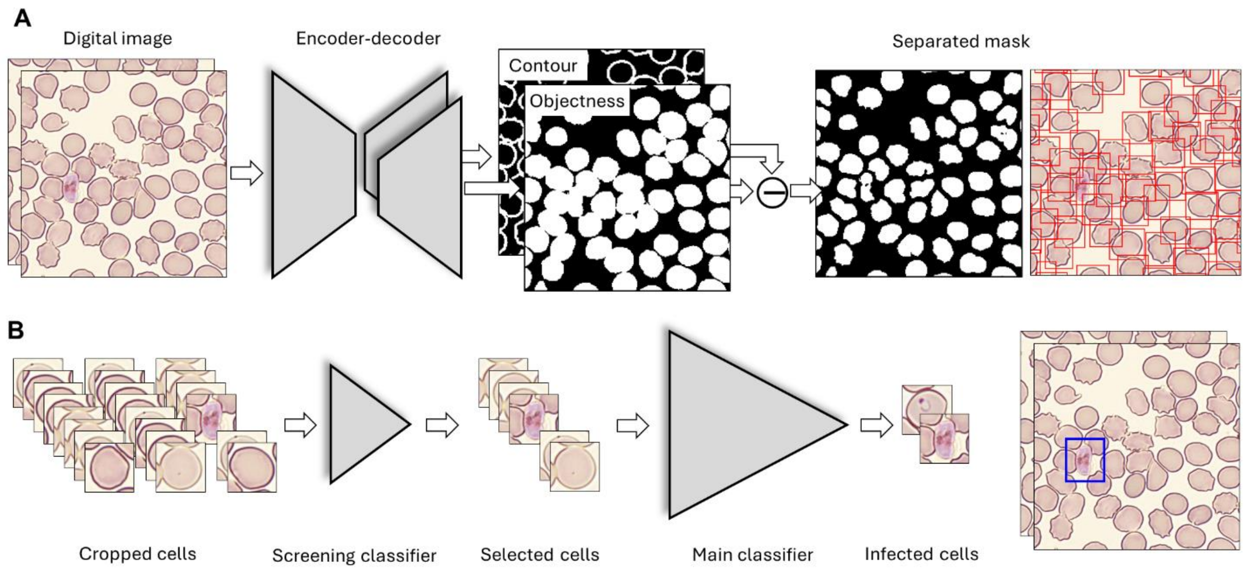


**Supplementary Figure 3. Schematic of the embedded deep-learning algorithm miLab$^{TM}$.** (A) The proposed segmentation network has a common encoder and two decoder branches. The network is trained to assign objectness value 1 when the corresponding pixel is on the foreground object, whereas the contour is trained to fire on the boundary of cells. The aggregated cells are separated by subtracting the contour image from the objectness image. (B) The cascaded neural network classifies the cropped RBCs.

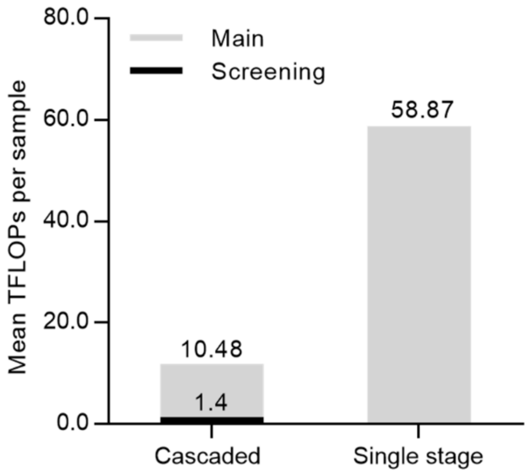


**Supplementary Figure 4. Efficiency of cascaded classifiers in the miLab™.** Although the cascaded classifiers have an additional screening classifier prior to the main classifier, the single stage classifier requires approximately 5 times more total computational cost to classify the RBCs.

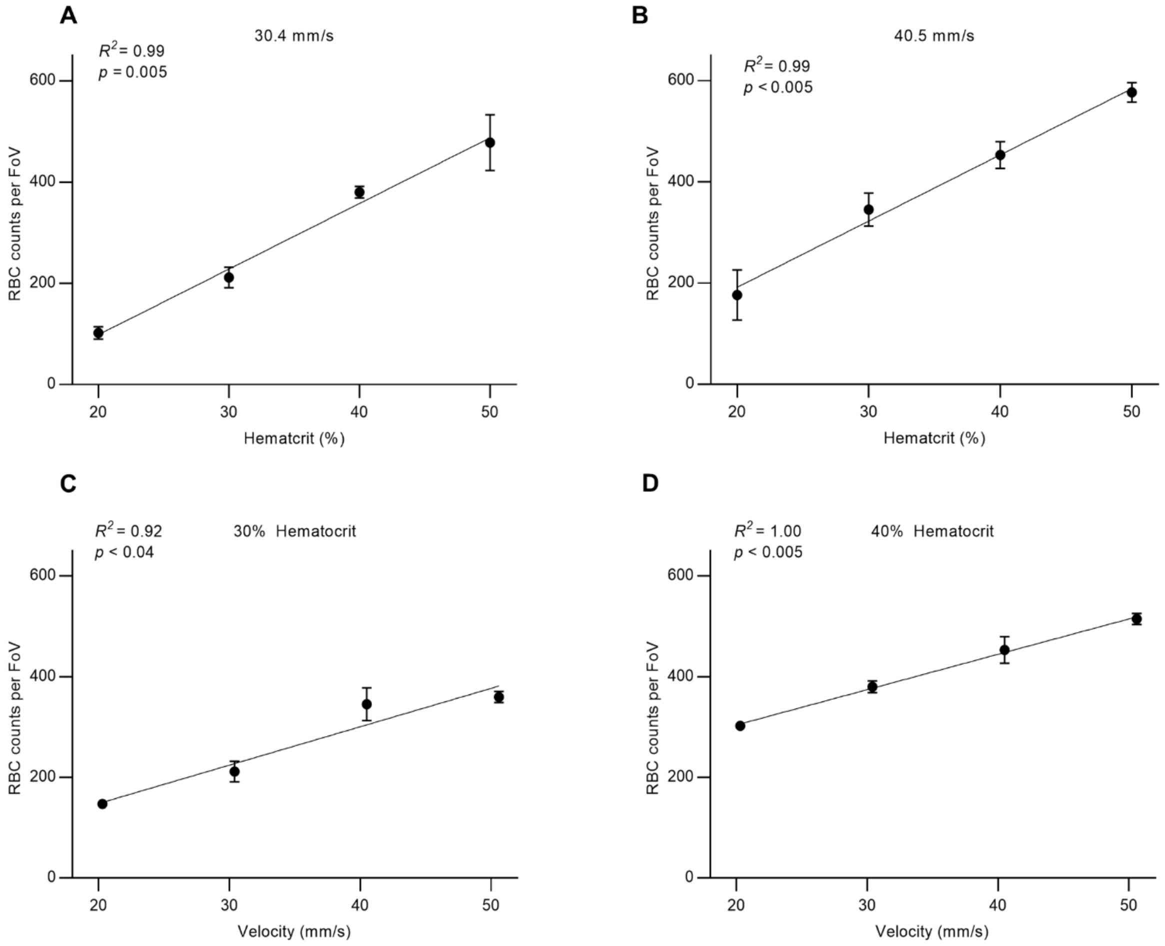


**Supplementary Figure 5. Optimization for the blood smear.** (A,B) Average counts of RBCs detected in a FoV depend on the smear velocity (20–50 mm/s). (C,D) Average counts of RBCs detected in a FoV depend on the hematocrit (20%–50%).

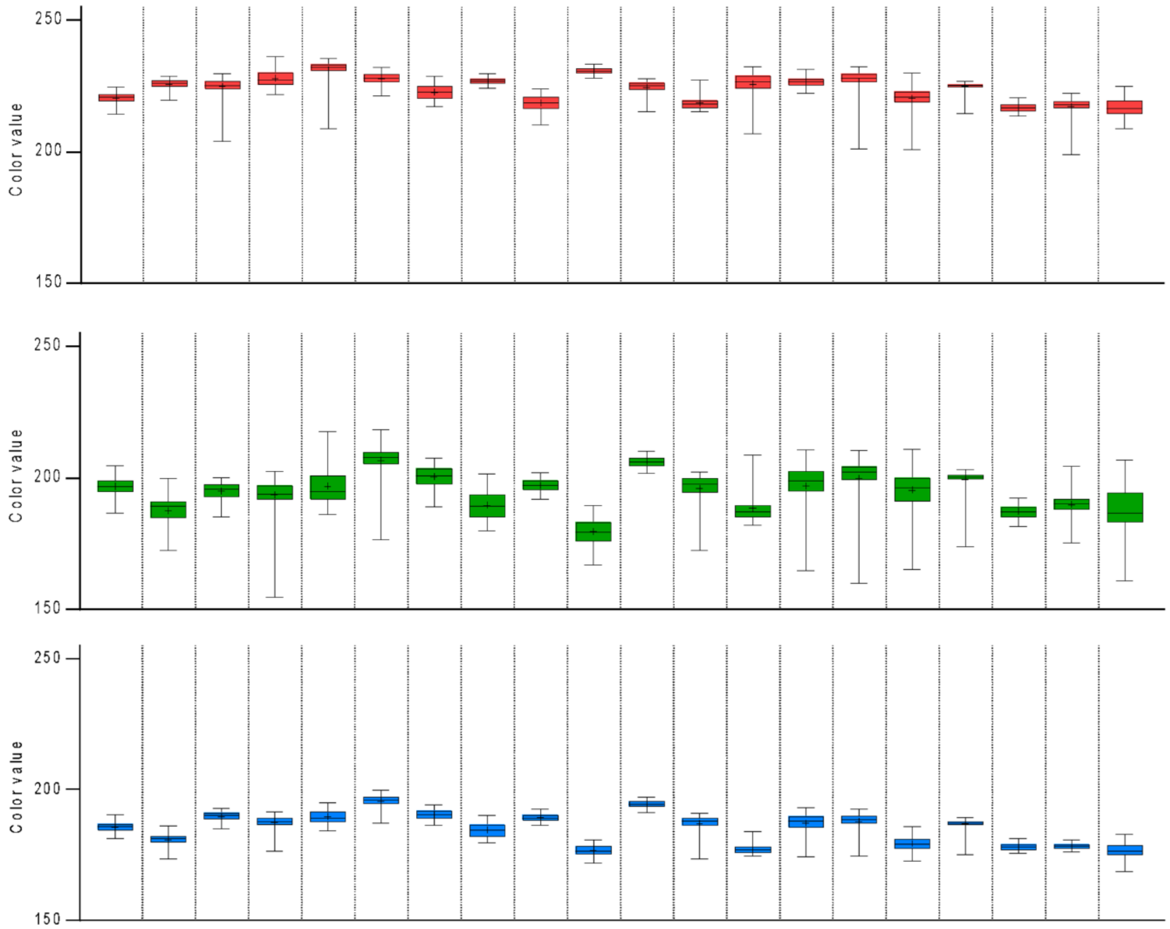


**Supplementary Figure 6. Color analysis for the verification of blood staining.** The color values of red, green, and blue in 20 clinical specimens are represented using box plots. The upper and lower ends of the error bars indicate the min and max value, respectively.

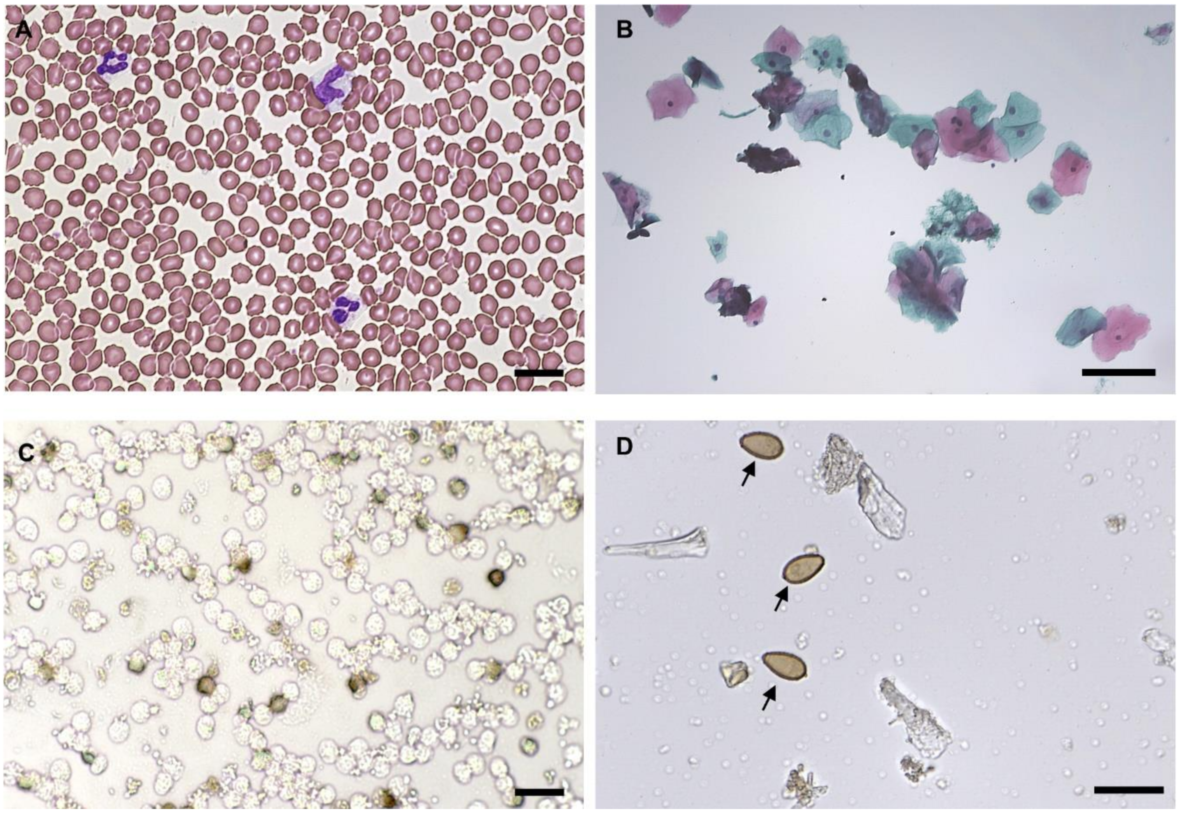


**Supplementary Figure 7. Application of miLab$^{TM}$ to diverse purpose.** Depending on the types of miLab™ cartridge, various dyes can be applied to the diverse clinical specimens such as (A) WBCs (neutrophil, monocyte), (B) Papanicolaou stained cells from cervix, (C) INF-gamma expressing peripheral blood mononuclear cells (PBMCs), and (D) Liver flukes (black arrow). Scale bars are (A) 20 μm, (B) 80 μm, (C) 20 μm, and (D) 40 μm, respectively.

### Supplementary Tables

### Supplementary Table 1. Clinical information (age) of patients.

| Age | Number | Percentage |
|---|---|---|
| **<5 years** | 58 | 10.45 |
| **5–18 years** | 156 | 28.11 |
| **>18 years** | 299 | 53.87 |
| **Unknown** | 42 | 7.57 |
| **Total** | 555 | 100.00 |
| **Age Average** | 21.58 | |
| **Age SD years** | 14.71 | |
| **Age Median (25-75 quartile)** | 20 | |

**Supplementary Table 2. Clinical information (sex) of patients.**

| Sex | Number | Percentage |
|---|---|---|
| **Female** | 336 | 60.54 |
| **Male** | 214 | 38.56 |
| **Unknown** | 5 | 0.90 |
| **Total** | 555 | 100.00 |
